# Systematic review and meta-analysis of the calibration of confidence judgments in individuals with schizophrenia spectrum disorders

**DOI:** 10.1101/2020.12.03.20243113

**Authors:** Martin Rouy, Pauline Saliou, Ladislas Nalborczyk, Michael Pereira, Paul Roux, Nathan Faivre

**Author notes:** **Corresponding author:** Martin Rouy, Laboratoire de Psychologie et Neurocognition, CNRS UMR 5105, UGA BSHM, 1251 Avenue Centrale, 38058 Grenoble Cedex 9. equal contribution (Paul Roux, Nathan Faivre). **Author Contributions:** MR, PS, MP, PR and NF developed the study concept and contributed to the study design. Data selection and extraction were performed by MR and PS. MR, LN and NF analyzed data. MR and NF drafted the paper; all authors provided critical revisions and approved the final version of the paper for submission. The authors declare no competing interests. **Data availability statement:** bibliographic data and analyses scripts are publicly available: https://gitlab.com/nfaivre/meta_analysis_scz_public.

## Abstract

**Background:** Metacognitive deficits are well documented in schizophrenia spectrum disorders as a decreased capacity to adjust confidence to first-order performance in a cognitive task. Because metacognitive ability directly depends on first-order performance, observed metacognitive deficits might be driven by lower first-order performance. We aimed to determine the extent to which individuals with schizophrenia experience specific deficits when producing confidence judgments and examined whether studies controlling for first-order performance found metacognitive deficits of smaller magnitude.

**Method:** Electronic databases were searched for studies published until April 24^th^ 2020. We conducted a Bayesian meta-analysis of 43 studies comparing the calibration of confidence in 1458 individuals with schizophrenia compared to 1337 matched controls. Group analyses and meta-regressions quantified how metacognitive deficits depended on task performance, cognitive domains, clinical severity, and antipsychotic dosage.

**Outcomes:** We found a global metacognitive deficit in schizophrenia (g = -0.57, 95% CrI [-0.71, -0.43]), which was driven by studies which did not equate first-order performance between groups (g = -0.64, 95% CrI [-0.77, -0.51]), and inconclusive among controlled-studies (g = -0.28, 95% CrI [-0.63, 0.07], BF_01_ = 1.3). Plus, the metacognitive deficit in non-controlled studies was correlated with first-order performance. No correlation was found between metacognitive deficit and clinical features of schizophrenia.

**Interpretation:** We provide evidence for the existence of a deficit in the calibration of confidence judgments in schizophrenia, which is inflated due to non-equated first-order performance. Thus, efforts should be made to develop experimental protocols accounting for lower first-order performance in schizophrenia.

## Introduction

Metacognition is the ability to monitor and control our own mental processes. Metacognitive deficits are thought to play an important role in schizophrenia spectrum disorders (hereafter: schizophrenia) (Hasson-Ohayon et al., 2018). These deficits are inferred both from subjective structured interviews (Semerari et al., 2003) and objective neuropsychological tasks (Koren et al., 2006), and have been linked to core features of schizophrenia including positive and negative symptoms (McLeod et al., 2014), lack of insight into illness (David et al., 2012), disorganisation (Vohs et al., 2014), functioning (Davies & Greenwood, 2020), and quality of life (Arnon-Ribenfeld et al., 2017).

Despite numerous studies, no meta-analysis has yet been conducted to examine metacognition in schizophrenia. Here we sought to conduct a systematic review and meta-analysis of neuropsychological measures of metacognitive performance in schizophrenia compared to matched healthy controls. From an experimental perspective, the gold standard to quantify metacognition is to assess how participants perform an experimental task (first-order task) and reflect on their own accuracy via confidence ratings (second-order task). Several studies employing this design have reported lower metacognitive performance in schizophrenia compared to healthy controls across different cognitive domains such as vision (Dietrichkeit et al., 2020; Jia et al., 2020; Moritz et al., 2014), audition (Gaweda & Moritz, 2019), emotion perception (Kother et al., 2012; Moritz et al., 2012; Pinkham et al., 2018), and memory (Berna et al., 2019; Mayer & Park, 2012; Moritz & Woodward, 2006). However, these results are mitigated by recent studies that failed to reveal such metacognitive deficits (Faivre et al., 2019; Powers et al., 2017; Wright et al., 2020). Noticeably these studies controlled for potential group differences in first-order performance, either at the design level through adaptive procedures (Levitt, 1971), or at the metric level through indices of metacognitive performance which are independent from first-order performance (Maniscalco & Lau, 2012). This is especially important in schizophrenia where cognitive impairments are well documented (Gopal & Variend, 2005; Heinrichs & Zakzanis, 1998) and associated with metacognitive deficits (Davies & Greenwood, 2020). Thus, a putative metacognitive deficit may be specific to second-order processing or merely inherited from a deficit at the first-order level. To determine whether schizophrenia involves specific deficits in the calibration of confidence, we conducted a systematic review followed by a Bayesian meta-analysis on a sample of 43 studies. We hypothesized that metacognitive deficits would be smaller in studies controlling for first-order performance. Following a pre-registered plan, we conducted additional subgroup analyses and meta-regressions to explore if metacognitive deficits vary across cognitive domains, severity of schizophrenia symptoms, and antipsychotic dosage.

## Methods

This meta-analysis followed the PRISMA recommendations (Moher et al., 2009). The protocol was registered on PROSPERO (CRD42020188614) on May 26^th^ 2020, before data extraction. Inclusion criteria followed the PICO framework.

- *Population*: individuals with schizophrenia or related disorders (schizoaffective, schizophreniform), as defined by standard diagnostic criteria (DSM-III, DSM-III-R, DSM-IV, DSM-IV-R, DSM-IV-TR, DSM 5, ICD-10).
- *Intervention* : a computerized or manual experimental task with self-reported confidence judgments as behavioral measures on a confidence scale with more than one trial.
- *Comparison* : healthy controls.
- *Outcome* : meta-performance defined as the strength of the relationship between first-order performance (accuracy on a neuropsychological task in perception, memory, executive functions, social cognition, and agency) and confidence in the first-order performance, repeated for each trial. Meta-performance indices included: meta-d’, M-Ratio, AUROC2, logistic regression, confidence gap, knowledge corruption index, gamma correlation (for details on these measures, see Fleming & Lau, 2014).

Search strategy: we retrieved English written preprints and peer-reviewed articles in three databases – Pubmed, Web of Science, Scopus – with the following query applied to the title, abstract and keywords:

*(schizophrenia OR schizophrenic OR schizo-affective OR schizoaffective) AND (confident OR confidence OR metacognition OR metacognitive OR “error awareness” OR “error monitoring”)*.

Search was performed on April 24^th^ 2020, and no new search before analysis was performed. This query could not identify one article (Powers et al., 2017) previously known by a co-author as it contained non-matching key-words and reported metacognitive performance in supplementary materials. It was manually included in the list of publications. Two authors (MR and PS) screened studies for inclusion in parallel, using Cadima (https://www.cadima.info; see supplementary information (SI) for details). For each study group, MR and PS extracted the following primary outcomes:

- whether the study controlled for first-order performance between groups (TRUE or FALSE)
- metacognitive performance indices (see above)
- first-order accuracy (% correct, d’)

Depending on the data available, either the mean and standard deviation, or raw statistics (t and F values) were extracted (SI). The following secondary outcomes were extracted:

- cognitive domain
- clinical characteristics including Positive and Negative Syndrome Scale scores (PANSS total, positive, and negative) and antipsychotic dosage (chlorpromazine equivalent).
- age (mean and standard deviation)
- sample size

All analyses were conducted in R. We used the brms package (Bürkner, 2017) based on the Stan framework (Carpenter et al., 2017) to fit a Bayesian meta-analytic multilevel model M1 with fixed and random effects as follows:

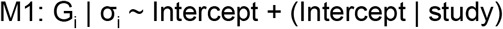

Where G_i_ denotes the Hedge’s g effect size of study i,σ _i_ denotes the standard error of the effect size from study i (SI). M1 estimated the overall effect-size of a difference in metacognitive performance between groups (the grand intercept of the model) while accounting for the between-study variability (random intercept per study; see SI for prior definition). To test the existence of a metacognitive deficit in schizophrenia (H1), we compared the estimations of M1 to the estimations of an alternative model M0 assuming that metacognitive deficit was inexistent (i.e., fixing the intercept at 0; H0).

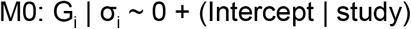

Results were interpreted based on the relative evidence toward H0 or H1 given by the Bayes factor (BF), and the statistics of the posterior distribution (mean and 95% credible interval, CrI).

Three measures of heterogeneity were computed: the Q-statistic (Card & Little, 2016), the Q-between statistic (Borenstein et al., 2010), and the I^2^ index for the percentage of the total variation due to between-studies variability (Higgins & Thompson, 2002). I^2^ values between 0 and 0.25 suggest small magnitudes of heterogeneity, 0.25 to 0.50 medium magnitudes, and > 0.50 large magnitudes. Exploratory subgroup analyses and meta-regressions were performed in case of significant Q-between and I^2^ above 25% (Sterne & Harbord, 2004). Namely, we assessed the metacognitive deficit amplitude across cognitive domains by fitting a model identical to M1 with the between-study variable “cognitive domains” (perception, memory, others) as an additional categorical covariate. We also explored the correlation between metacognitive performance among patients and continuous variables by adding standardized (z-scores) PANSS scores and chlorpromazine equivalent as meta-regressors to M1.

To quantify the risk of bias in individual studies, we assessed whether our selection contained extreme effect size values via a leave-one-out sensitivity analysis (SI). We also assessed the risk of bias according to the Newcastle-Ottawa Scale (NOS) adapted for case-control studies (SI). Publication bias was assessed using a funnel plot of observed outcomes against corresponding standard errors (Sterne & Harbord, 2004). The distribution of p-values was analyzed using the R package dmetar (Harrer et al., 2019) to examine whether some of the studies were subject to p-hacking (p-curve: Simonsohn et al., 2014).

## Results

Our search retrieved 13933 records, 7886 after duplicates removal. 7745 records were excluded after title and abstract screening (Fig.1). Another 99 articles were excluded on the basis of full-text screening, resulting in a selection of 42 articles. One study was excluded because of a strongly deviant outcome identified via a leave-one-out analysis performed on the metacognitive deficit effect-size (SI). Among the 41 remaining articles, two were split into two independent studies as they involved different populations (young versus old: Gaweda et al, 2015; hallucination-prone versus non-hallucination-prone: Powers et al., 2017). The final selection consisted of 43 studies, with a total population of 2795 participants (1458 patients) (Table 1).

**Figure 1:**
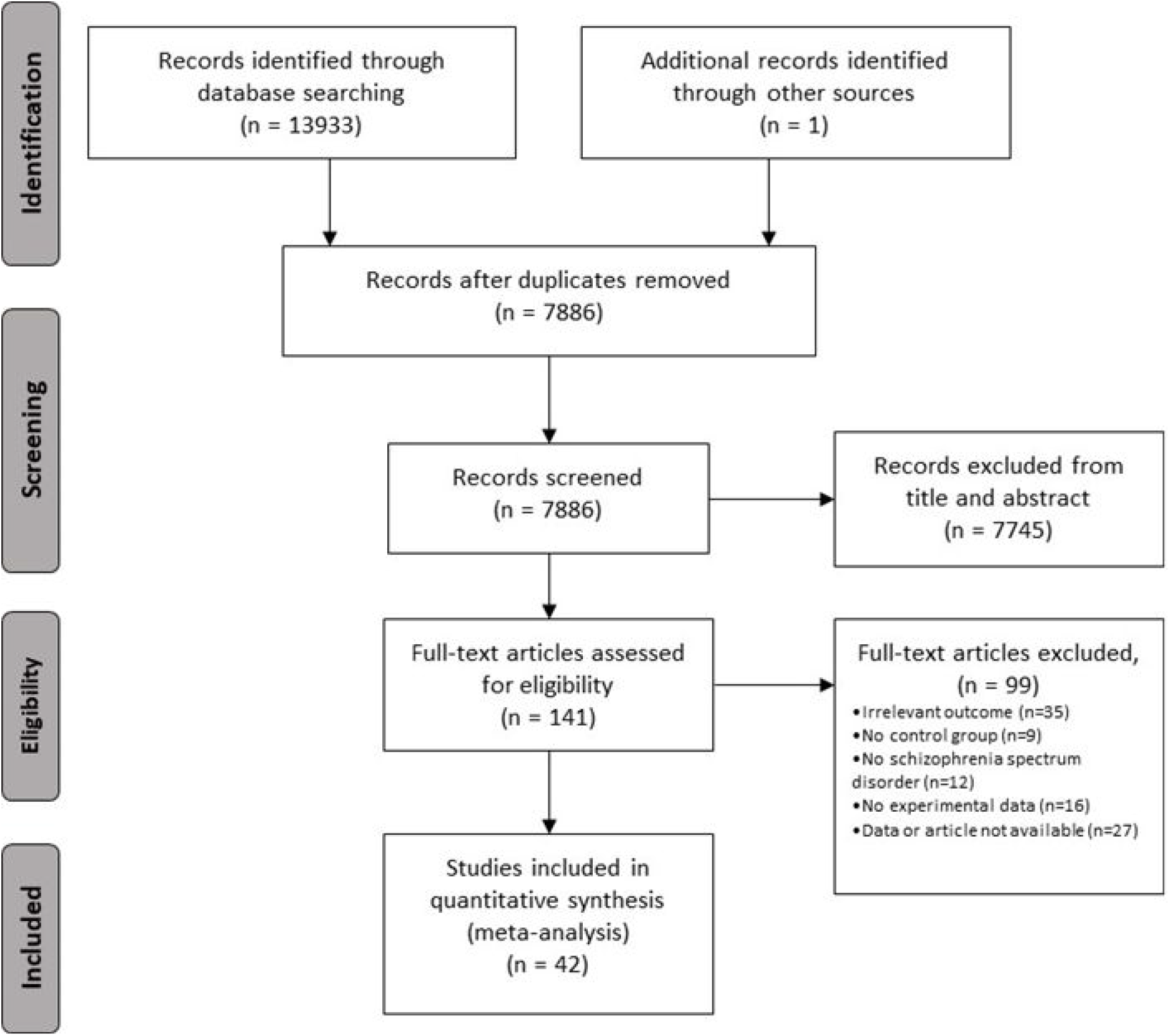
Flow diagram of the selection process.

**Table 1:**
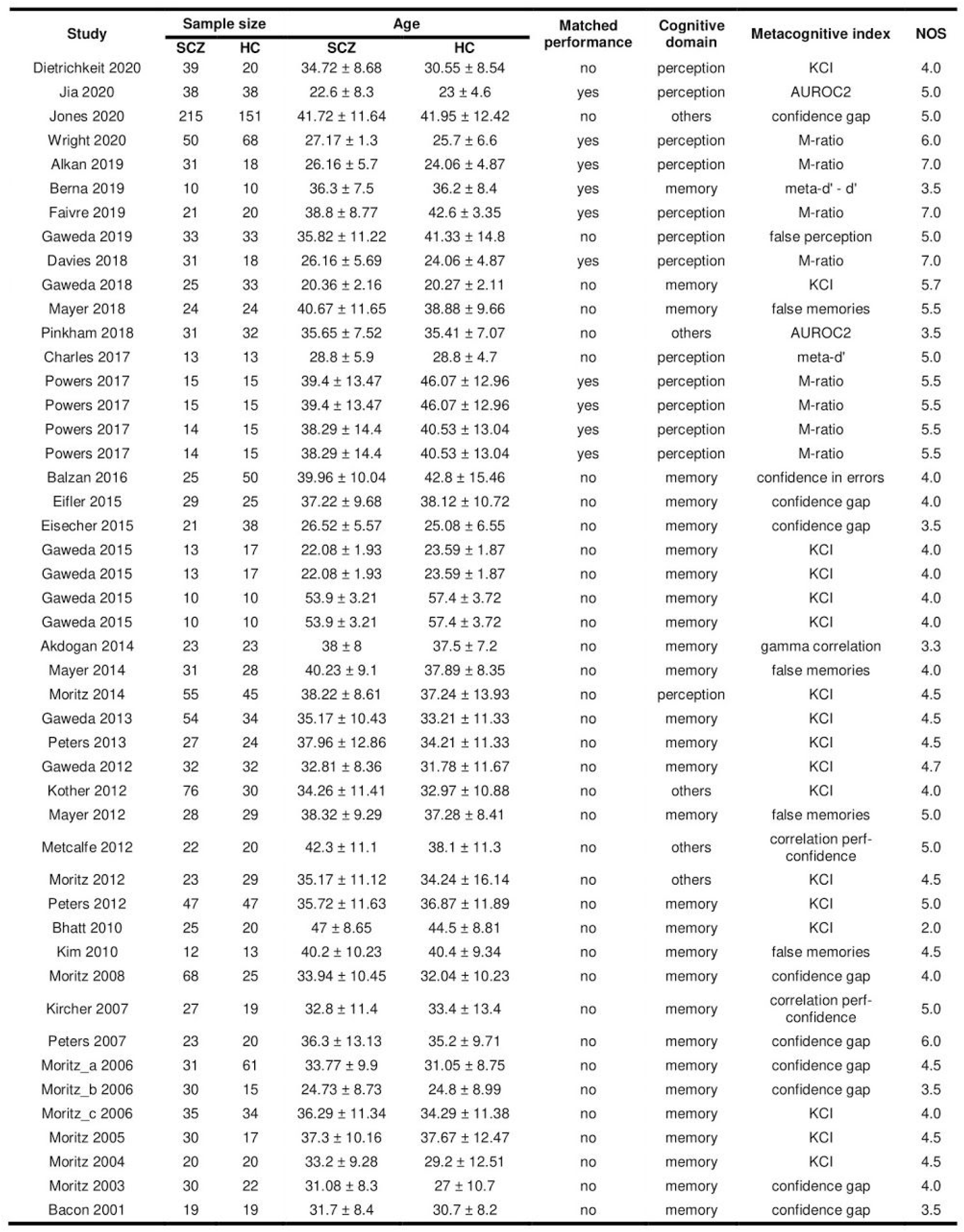
study characteristics. KCI: knowledge corruption index; AUROC2: area under the type 2 receiver operating characteristic curve.

| Study | Sample size |  | Age |  | Matched performance | Cognitive domain | Metacognitive index | NOS |
| --- | --- | --- | --- | --- | --- | --- | --- | --- |
|  | SCZ | HC | SCZ | HC |  |  |  |  |
| Dietrichkeit 2020 | 39 | 20 | 34.72 ± 8.68 | 30.55 ± 8.54 | no | perception | KCI | 4.0 |
| Jia 2020 | 38 | 38 | 22.6 ± 8.3 | 23 ± 4.6 | yes | perception | AUROC2 | 5.0 |
| Jones 2020 | 215 | 151 | 41.72 ± 11.64 | 41.95 ± 12.42 | no | others | confidence gap | 5.0 |
| Wright 2020 | 50 | 68 | 27.17 ± 1.3 | 25.7 ± 6.6 | yes | perception | M-ratio | 6.0 |
| Alkan 2019 | 31 | 18 | 26.16 ± 5.7 | 24.06 ± 4.87 | yes | perception | M-ratio | 7.0 |
| Berna 2019 | 10 | 10 | 36.3 ± 7.5 | 36.2 ± 8.4 | yes | memory | meta-d' - d' | 3.5 |
| Faivre 2019 | 21 | 20 | 38.8 ± 8.77 | 42.6 ± 3.35 | yes | perception | M-ratio | 7.0 |
| Gaweda 2019 | 33 | 33 | 35.82 ± 11.22 | 41.33 ± 14.8 | no | perception | false perception | 5.0 |
| Davies 2018 | 31 | 18 | 26.16 ± 5.69 | 24.06 ± 4.87 | yes | perception | M-ratio | 7.0 |
| Gaweda 2018 | 25 | 33 | 20.36 ± 2.16 | 20.27 ± 2.11 | no | memory | KCI | 5.7 |
| Mayer 2018 | 24 | 24 | 40.67 ± 11.65 | 38.88 ± 9.66 | no | memory | false memories | 5.5 |
| Pinkham 2018 | 31 | 32 | 35.65 ± 7.52 | 35.41 ± 7.07 | no | others | AUROC2 | 3.5 |
| Charles 2017 | 13 | 13 | 28.8 ± 5.9 | 28.8 ± 4.7 | no | perception | meta-d' | 5.0 |
| Powers 2017 | 15 | 15 | 39.4 ± 13.47 | 46.07 ± 12.96 | yes | perception | M-ratio | 5.5 |
| Powers 2017 | 15 | 15 | 39.4 ± 13.47 | 46.07 ± 12.96 | yes | perception | M-ratio | 5.5 |
| Powers 2017 | 14 | 15 | 38.29 ± 14.4 | 40.53 ± 13.04 | yes | perception | M-ratio | 5.5 |
| Powers 2017 | 14 | 15 | 38.29 ± 14.4 | 40.53 ± 13.04 | yes | perception | M-ratio | 5.5 |
| Balzan 2016 | 25 | 50 | 39.96 ± 10.04 | 42.8 ± 15.46 | no | memory | confidence in errors | 4.0 |
| Eifler 2015 | 29 | 25 | 37.22 ± 9.68 | 38.12 ± 10.72 | no | memory | confidence gap | 4.0 |
| Eisecher 2015 | 21 | 38 | 26.52 ± 5.57 | 25.08 ± 6.55 | no | memory | confidence gap | 3.5 |
| Gaweda 2015 | 13 | 17 | 22.08 ± 1.93 | 23.59 ± 1.87 | no | memory | KCI | 4.0 |
| Gaweda 2015 | 13 | 17 | 22.08 ± 1.93 | 23.59 ± 1.87 | no | memory | KCI | 4.0 |
| Gaweda 2015 | 10 | 10 | 53.9 ± 3.21 | 57.4 ± 3.72 | no | memory | KCI | 4.0 |
| Gaweda 2015 | 10 | 10 | 53.9 ± 3.21 | 57.4 ± 3.72 | no | memory | KCI | 4.0 |
| Akdogan 2014 | 23 | 23 | 38 ± 8 | 37.5 ± 7.2 | no | memory | gamma correlation | 3.3 |
| Mayer 2014 | 31 | 28 | 40.23 ± 9.1 | 37.89 ± 8.35 | no | memory | false memories | 4.0 |
| Moritz 2014 | 55 | 45 | 38.22 ± 8.61 | 37.24 ± 13.93 | no | perception | KCI | 4.5 |
| Gaweda 2013 | 54 | 34 | 35.17 ± 10.43 | 33.21 ± 11.33 | no | memory | KCI | 4.5 |
| Peters 2013 | 27 | 24 | 37.96 ± 12.86 | 34.21 ± 11.33 | no | memory | KCI | 4.5 |
| Gaweda 2012 | 32 | 32 | 32.81 ± 8.36 | 31.78 ± 11.67 | no | memory | KCI | 4.7 |
| Kother 2012 | 76 | 30 | 34.26 ± 11.41 | 32.97 ± 10.88 | no | others | KCI | 4.0 |
| Mayer 2012 | 28 | 29 | 38.32 ± 9.29 | 37.28 ± 8.41 | no | memory | false memories | 5.0 |
| Metcalfe 2012 | 22 | 20 | 42.3 ± 11.1 | 38.1 ± 11.3 | no | others | correlation perf-confidence | 5.0 |
| Moritz 2012 | 23 | 29 | 35.17 ± 11.12 | 34.24 ± 16.14 | no | others | KCI | 4.5 |
| Peters 2012 | 47 | 47 | 35.72 ± 11.63 | 36.87 ± 11.89 | no | memory | KCI | 5.0 |
| Bhatt 2010 | 25 | 20 | 47 ± 8.65 | 44.5 ± 8.81 | no | memory | KCI | 2.0 |
| Kim 2010 | 12 | 13 | 40.2 ± 10.23 | 40.4 ± 9.34 | no | memory | false memories | 4.5 |
| Moritz 2008 | 68 | 25 | 33.94 ± 10.45 | 32.04 ± 10.23 | no | memory | confidence gap | 4.0 |
| Kircher 2007 | 27 | 19 | 32.8 ± 11.4 | 33.4 ± 13.4 | no | memory | correlation perf-confidence | 5.0 |
| Peters 2007 | 23 | 20 | 36.3 ± 13.13 | 35.2 ± 9.71 | no | memory | confidence gap | 6.0 |
| Moritz_a 2006 | 31 | 61 | 33.77 ± 9.9 | 31.05 ± 8.75 | no | memory | confidence gap | 4.5 |
| Moritz_b 2006 | 30 | 15 | 24.73 ± 8.73 | 24.8 ± 8.99 | no | memory | confidence gap | 3.5 |
| Moritz_c 2006 | 35 | 34 | 36.29 ± 11.34 | 34.29 ± 11.38 | no | memory | KCI | 4.0 |
| Moritz 2005 | 30 | 17 | 37.3 ± 10.16 | 37.67 ± 12.47 | no | memory | KCI | 4.5 |
| Moritz 2004 | 20 | 20 | 33.2 ± 9.28 | 29.2 ± 12.51 | no | memory | KCI | 4.5 |
| Moritz 2003 | 30 | 22 | 31.08 ± 8.3 | 27 ± 10.7 | no | memory | confidence gap | 4.0 |
| Bacon 2001 | 19 | 19 | 31.7 ± 8.4 | 30.7 ± 8.2 | no | memory | confidence gap | 3.5 |

Our selection included 11 perception (auditory and visual), 27 memory, 4 social cognition, and 1 agency studies. Because of their low number, social cognition and agency studies were regrouped into a generic category termed “others”.

The meta-analytic model M1 revealed lower metacognitive performance in the schizophrenia vs. control groups with an effect size g = -0.57, 95% CrI [-0.71, -0.43] (Fig.2). Comparison against the null hypothesis (i.e., absence of a metacognitive deficit in schizophrenia modelled by M0) resulted in a Bayes factor favoring the alternative hypothesis BF_10_ = 78.58 × 10^6^, indicating extremely strong evidence in favor of a metacognitive deficit in schizophrenia. Of note, this pattern of results was robust to prior variations (SI).

**Figure 2:**
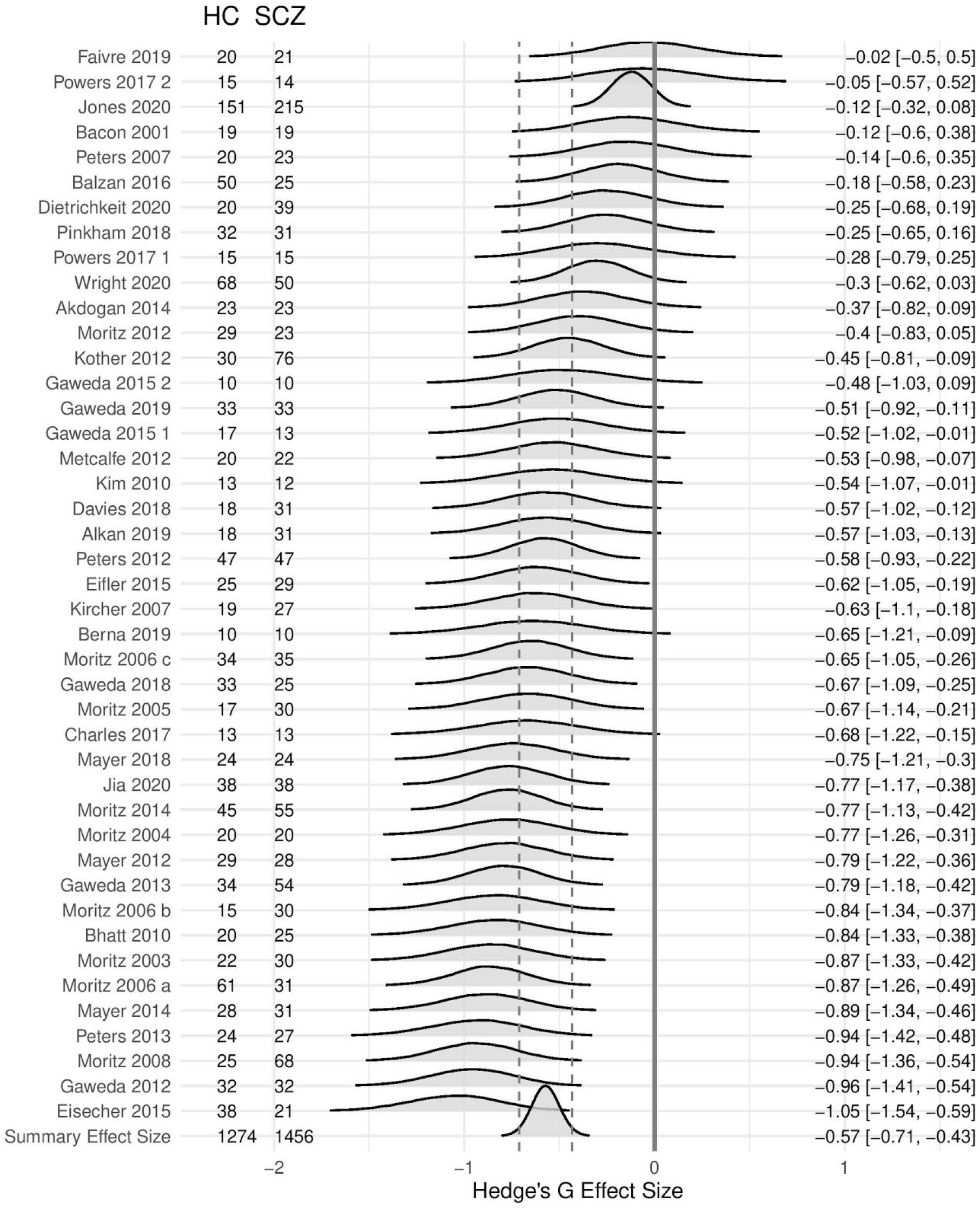
Forest plot of the metacognitive deficit in schizophrenia. Left: Authors with publication year and sample sizes; Middle: posterior distribution of the effect size; Right: mean and 95% CrI of the posterior distribution. The summary effect size is displayed on the last row: the solid vertical grey line is centred on zero (i.e., equivalent metacognitive performance between groups), and the dashed vertical lines depict the boundaries of the 95% CrI.

Heterogeneity analyses produced a significant Q-statistic (124.6, df = 42, p < .001) and a high amount of heterogeneity (I^2^ statistic 0.66, 95% CI [0.53, 0.75]), suggesting moderator analyses were appropriate. Because metacognitive performance is known to depend on first-order performance (Maniscalco & Lau, 2012), and because the latter differed between groups (g = -0.64, 95% CrI [-0.77, -0.51], BF_10_ = 2.11 × 10^10^), we sought to assess whether metacognitive deficits could stem from cognitive impairments that are well documented in schizophrenia (Gopal & Variend, 2005; Heinrichs & Zakzanis, 1998). Distinguishing studies controlling for first-order performance (N = 8) from those which did not (N = 35) revealed a significant moderation effect (Q-between = 5.89, df = 1, p = 0.015). Thus, we assessed the influence of performance-matching with a model identical to M1 including performance-matching as an additional binary predictor. The sub-group of non-controlled studies had an overall metacognitive deficit of magnitude g = -0.63, 95% CrI [-0.79, -0.49], which was reduced to g = -0.28, 95% CrI [-0.63, 0.07] in the sub-group of controlled studies (Fig.3A). Accordingly, the evidence ratio supporting our directional hypothesis that controlling for first-order performance decreases the magnitude of the metacognitive deficit was very strong (BF_10_ = 41) (Fig.3B). Comparison against the null hypothesis among controlled studies revealed inconclusive evidence in favor of a metacognitive deficit in schizophrenia (BF_01_ = 1.3). Finally, a positive correlation between cognitive and metacognitive deficits was found among non-controlled studies (SI). Sub-group analyses reduced heterogeneity which however remained significant (SI).

**Figure 3:**
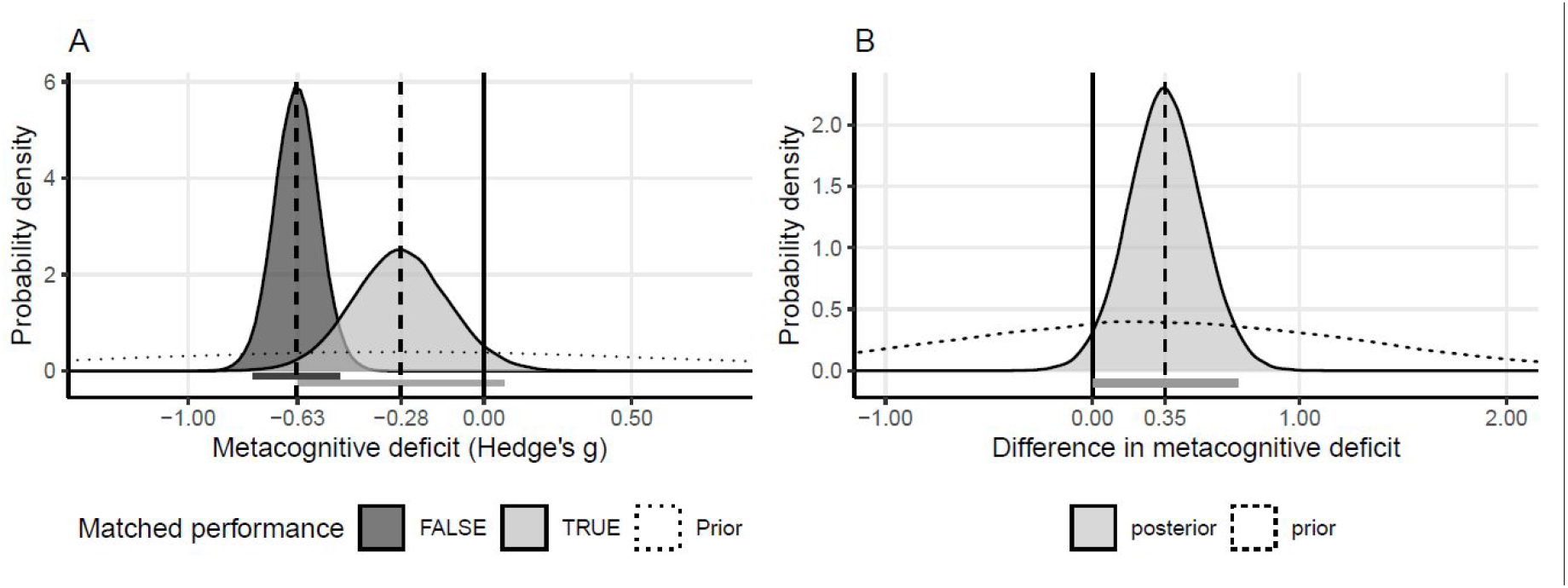
A: Posterior distributions of the metacognitive deficit. Dark gray: non-controlled first-order performance (n=35), Light gray: controlled first-order performance (n=8). B: Posterior distribution of the difference in effect size between studies which did or did not control for first-order performance. In both panels, dotted lines represent the prior distributions, vertical dashed lines the mean posterior values, and the horizontal bars the 95% CrI.

Next, in line with our pre-registered analysis plan and a significant moderation effect of cognitive domains (Qbetween = 38.2, df = 2, p < .001), we assessed how metacognitive deficits varied across cognitive domains (i.e., perception, memory, others). A subgroup analysis revealed the largest metacognitive deficit among memory studies, compared to perception and others. Mean value of the metacognitive deficit in the memory domain (g = -0.74, 95 % CrI [-0.89, -0.58], BF_10_ = 7.74 × 10^156^) was twice higher than in the perception domain (g = -0.35, 95 % CrI [-0.63, -0.07], BF_10_ = 4.12), and three times higher than in other domains (g = -0.26, 95 % CrI [-0.62, 0.09], BF_10_ = 0.39; see Fig.4 and SI). Sub-group analyses reduced heterogeneity which however remained significant (SI).

Finally, we performed further meta-regressions to explore how metacognitive deficits co-varied with the severity of positive and negative symptoms (PANSS equivalent scores) and antipsychotic dosage (chlorpromazine equivalent). Contrary to what we had predicted, none of these meta-regressions revealed conclusive evidence (SI).

**Figure 4:**
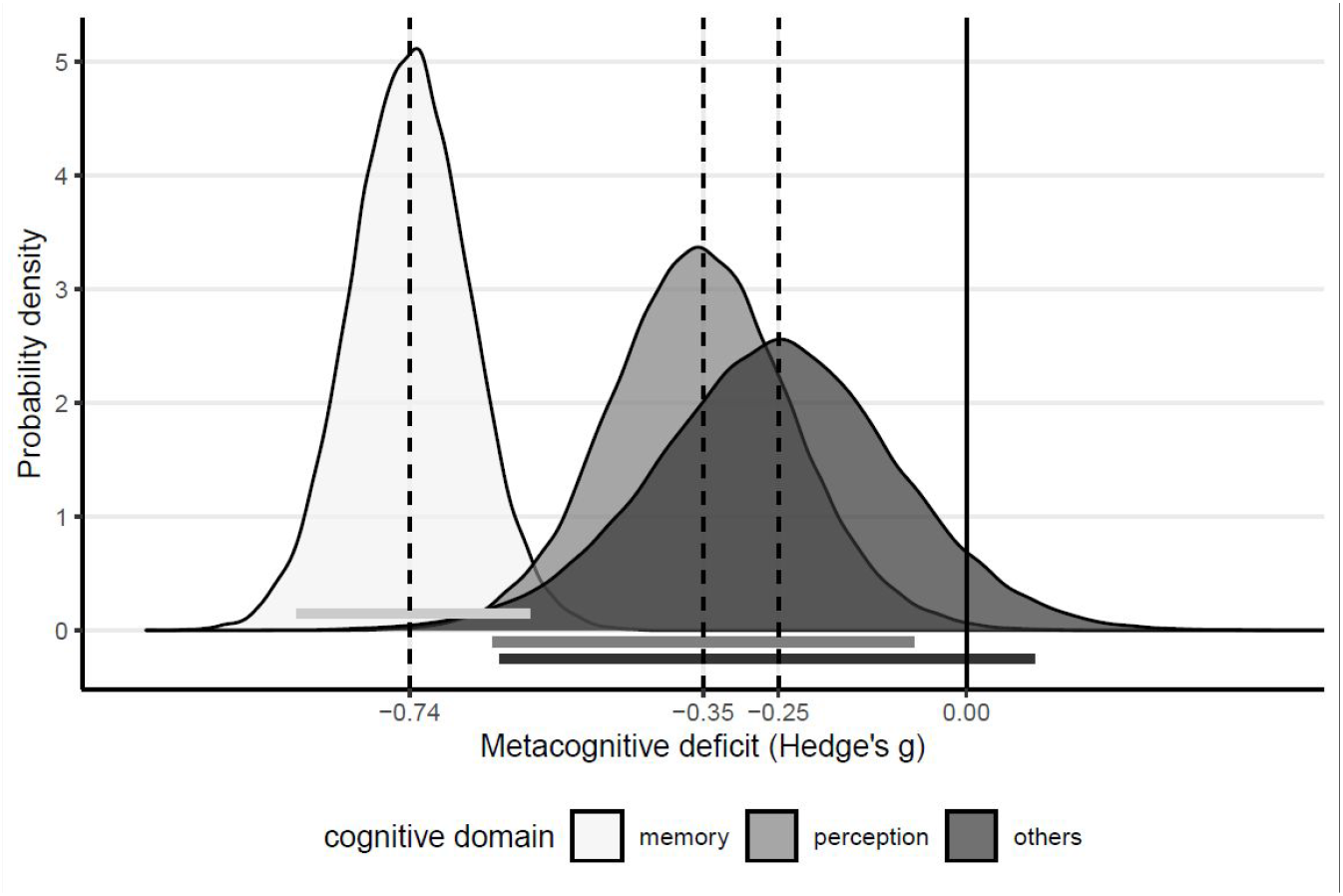
Posterior distributions of the metacognitive deficit (Hedge’s g effect size) according to each cognitive domain. The vertical dashed lines represent mean values and the horizontal bars the 95% CrI.

### Risk of bias in selected studies

A quality evaluation using the Newcastle-Ottawa Scale suggested that about half the studies had a relatively high risk of bias with scores < 5/9 (SI and Luchini et al., 2017). The shape of the funnel plot revealed no asymmetry (Egger’s test: z = - 0.46, p = 0.65; Fig.5A and SI), suggesting no clear publication bias. Plus, testing the right-skewness of the P-curve (Fig.5B) with Stouffer’s method revealed that both the half (p’s < 0.025) and full p-curves (p’s < 0.05) were right-skewed with p < .001, suggesting that our study sample was not contaminated by p-hacking.

**Figure 5:**
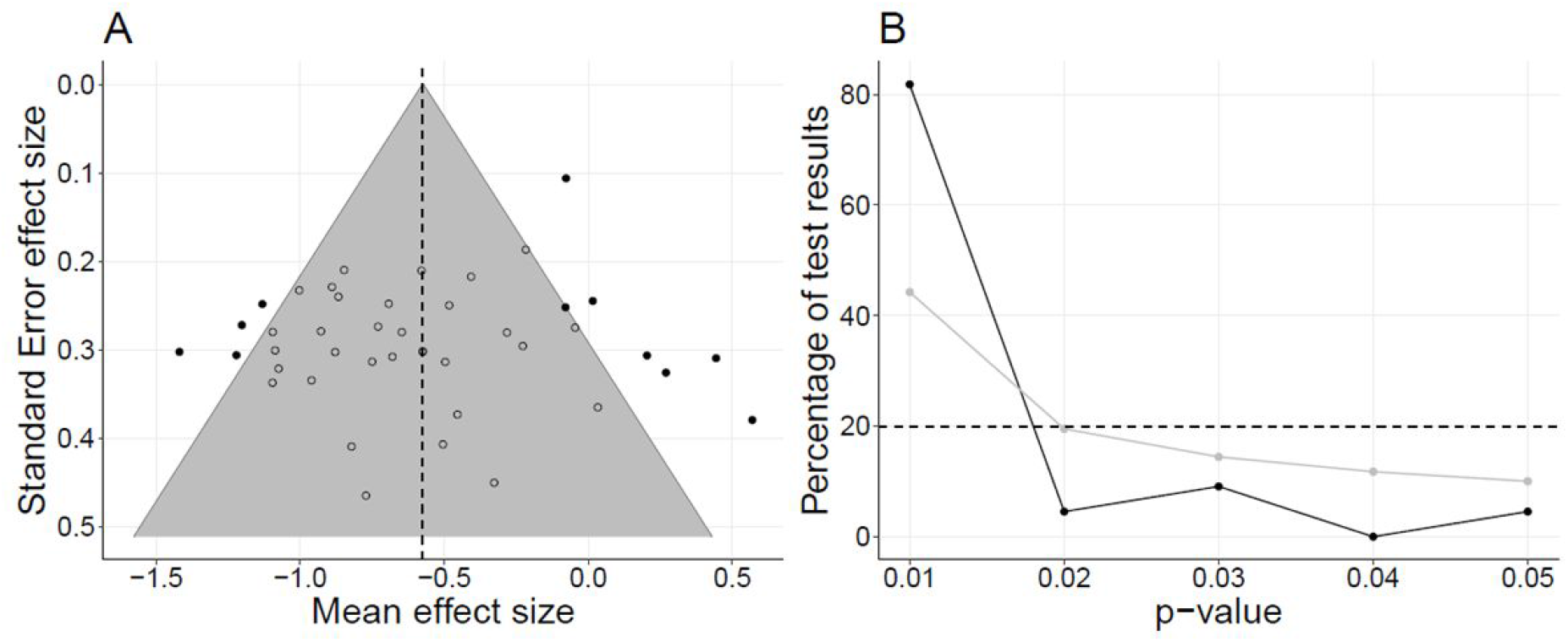
A: Funnel plot centered on the overall effect size. The vertical dashed line represents the global metacognitive deficit. The gray area represents the 95% CI of the overall effect size. Each dot represents a study, full dots represent outliers. B: Observed p-curve (black) and theoretical p-curve expected for low-powered (33%) studies (gray). Horizontal dashed line: Expected uniform distribution for null effects.

## Discussion

The present meta-analysis based on 43 studies and 2795 individuals aimed at synthesizing the literature on the calibration of confidence judgments among individuals with schizophrenia. At first sight, our findings confirmed a deficit in the calibration of confidence judgments in schizophrenia, but with high heterogeneity. The effect was of medium magnitude, which is smaller than the large effects reported in prior meta-analyses regarding cognitive impairments (Schaefer et al., 2013). The leave-one-out sensitivity analysis confirmed this effect was robust to outliers. We found several sources for heterogeneity that we describe hereafter.

Because patients’ first-order cognitive deficits risked to artificially inflate metacognitive deficits (Galvin et al., 2003), we hypothesized that metacognitive deficits would be reduced in studies equating first-order performance between groups. Results indicated strong evidence in favor of our hypothesis, as metacognitive deficits were twice smaller in studies controlling for first-order performance, most of them concerning the perceptual domain. In this subset of studies, assessing the presence of a metacognitive deficit revealed inconclusive evidence. By contrast, a correlation between cognitive and metacognitive deficits was found among non-controlled studies, indicating that first-order performance is a critical moderator of metacognition which should be controlled for when assessing metacognitive deficits in schizophrenia. We also explored possible differences in metacognitive deficits across cognitive domains (perception, memory, others), and found the most prominent deficits among memory studies. As such, this result is not sufficient to confirm the presence of a specific meta-memory deficit in schizophrenia, as all the memory studies but one did not control for differences in first-order performance between groups. Given that the magnitude of the meta-memory deficit we found is lower than the one of episodic verbal memory (range between -1.53 and -1.11 SD) (Gopal & Variend, 2005; Heinrichs & Zakzanis, 1998; Schaefer et al., 2013), arbitrating between the existence of a specific meta-memory deficit or the side effect of a non-controlled first-order factor will require the development of more robust experimental protocols. Of note, this meta-analysis did not examine the literature based on judgments of learning or feeling of knowing, which may reveal different patterns of results (Souchay et al., 2006).

Despite moderation analyses, heterogeneity remained high even after clustering studies according to performance matching and cognitive domains. This heterogeneity may be explained by the different diagnoses included in our selection of studies. The category of first episode of psychosis may be particularly problematic, as it included variable diagnoses (mania with psychosis, bipolar disorder with psychosis, depression with psychosis, delusional disorder, substance-induced psychotic disorder, psychosis not otherwise specified, acute and transient psychotic disorder, brief psychotic disorder). Heterogeneity may also come from the use of idiosyncratic first-order tasks (e.g., memory performance was quantified using recognition, source memory and spatial delayed response tasks) and confidence scales (e.g., ordinal vs. continuous scales, full vs. half scales, etc.). With this in mind, it will be important to use more systematic paradigms in the future.

Additional analyses evaluating how metacognitive deficits varied as a function of clinical scores (PANSS total, positive, negative) and antipsychotic dosage (chlorpromazine equivalent) revealed inconclusive evidence for correlation in each case. As we had no access to individual data, correlations were based on summary statistics extracted from each experimental group, which is clearly suboptimal. As with all meta-analyses, our findings are shaped and limited by selection and analytical methods, and the information made available to researchers in the studies selected for review. The scarcity of data prevented us from running planned analyses regarding the link between metacognitive performance and clinical/cognitive insight. Establishing this link is of crucial importance to validate confidence calibration as a valid empirical construct for clinical practice, and to refine current strategies to improve insight in schizophrenia. We encourage authors to share anonymized individual data similar to what is done for healthy controls (Rahnev et al., 2020) on a dedicated repository (https://osf.io/cfm5d/).

Our findings point to a number of areas for future research. First, few studies included in this meta-analysis measured mood, despite it being an important determinant of metacognition (Lin et al., 2019), with a bias toward underconfidence in depression (Hoven et al., 2019). No study included in this meta-analysis focused on the metacognition of executive function. Further studies are needed because meta-executive functions have been linked with attenuated psychosis syndrome (Koren et al., 2019). Further studies should also investigate whether the calibration of confidence judgments is associated with insight, relapse and psychosocial functioning before using it in clinical settings.

## Conclusion

This is the first meta-analysis to examine the deficit in the calibration of confidence judgments in schizophrenia. Our results show that this deficit is inflated due to non-equated first-order performance, and varies across cognitive domains. Importantly, metacognitive deficits may also be overestimated in other psychiatric and neurological conditions involving cognitive impairments. Efforts should be made to develop experimental protocols accounting for lower first-order performance in schizophrenia before including calibration of confidence judgment as a cognitive dimension in neuropsychological batteries for clinical applications.

## Data Availability

bibliographic data and analyses scripts are publicly available.

https://gitlab.com/nfaivre/meta_analysis_scz_public

## Acknowledgments

NF has received funding from the European Research Council (ERC) under the European Union’s Horizon 2020 research and innovation programme (Grant agreement No. 803122). We thank Mathieu Roger for his help at the beginning of this project. We thank Samuele Cortese and Clément Dondé for their valuable advice.

