## Supplementary information for "Systematic review and meta-analysis of the calibration of confidence judgments in individuals with schizophrenia spectrum disorders"

### Supplementary methods

Search strategy: In Scopus, we retrieved results from both “documents” and “secondary documents” tabs.

Data selection: to ensure mutual agreement, MR and PS iteratively performed consistency checks on 20 randomly selected studies prior to the selection process, until they reached 90% of agreement. The selection process consisted in the application of the inclusion criteria from the PICO scheme following a two-step process, first on the basis of title and abstract, and then on full-text. The raters were blind to each other’s decisions. The first step resulted in a total of 192 disagreements, which were resolved on the basis of information from the abstracts. The raters commented and double-checked each conflicting judgement until an agreement was reached. The same strategy was used for resolving the disagreements in the full-text selection.

Data extraction: when scores of interest were reported separately across variables irrelevant for our purpose, we computed weighted means and pooled standard deviations. In studies reporting several experiments from the same sample, we computed mean scores and pooled standard deviations, resulting in one score for each variable of interest per study. On the contrary, studies reporting scores from different population samples were considered as if they were independent experiments.

Clinical scores: Brief Psychiatric Rating Scale (BPRS), Scale for the Assessment of Positive Symptoms (SAPS) and Scale for the Assessment of Negative Symptoms (SANS) scores were transformed into PANSS equivalent scores according to the recommended formulae (Leucht et al., 2013; van Erp et al., 2014). We had further pre-registered the extraction of confidence bias (mean confidence), depression, insight, psychosocial functioning scores, and brief psychosis episode, but did not proceed due to too few studies reporting them (N = 9, N = 4, N = 6, N = 5, and N = 7, respectively).

Formulation of the meta-analytic model:

Let *ES* be the observed effect sizes:

$ES_i \sim \text{Normal}(\mu_i, \sigma_i)$

$\mu_i = \alpha + \alpha_{study[i]}$

$\alpha \sim \text{Normal}(-0.3, 1)$

$\alpha_{study[i]} \sim \text{Normal}(0, \tau)$

$\tau \sim \text{Half-Cauchy}(0.1)$

Where $\mu_i$ indicates the effect size of study *i*, and $\sigma_i^2$ is the known variance of the effect in study *i*.

$\alpha$ is the intercept parameter of the model (the average effect size in the population). We chose a random-effect model rather than a fixed-effect model as the distributions of effect sizes are expected to be heterogeneous for metacognition in schizophrenia. Because metacognitive deficit in schizophrenia is commonly described, we specified a mildly informative prior corresponding to a metacognitive deficit with small to medium effect-size, and $\tau^2$ is the between-studies variance, provided with a mildly informative prior. Because studies with multiple experiments were rare (4 in total), we could not add experiment in addition to study as a random factor in our model.

Computation of the effect sizes (Hedge' g):

Effect sizes were computed as Hedge's G with the R package esc (Lüdecke, 2018) using the procedure given in Borenstein et al. (2010) as follows:

$$g = J \times \frac{mean_p - mean_c}{sd_{pooled}}$$

where $mean_p$ is the average meta-performance reported for the patient group of size $n_p$, and $mean_c$ is the average meta-performance reported for the control group of size $n_c$.

$sd_{pooled}$ is the pooled standard deviation within both groups:

$$sd_{pooled} = \sqrt{\frac{(n_p - 1)sd_p^2 + (n_c - 1)sd_c^2}{n_p + n_c - 2}}$$

with $sd_p$ the standard deviation of the meta-performance reported for the patient group, and $sd_c$ the standard deviation for the control group.

The correction factor J to achieve an unbiased estimator is defined as:

$$J = 1 - \frac{3}{4\left(n_p + n_c - 2\right) - 1}$$

Finally, the variance of g is defined as:

$$var(g) = J^2.\left[\frac{n_p + n_c}{n_p n_c} + \frac{1}{2\left(n_p + n_c\right)}\left(\frac{mean_p - mean_c}{sd_{pooled}}\right)^2\right]$$

Hypothesis testing: we assessed the relative evidence toward H0 (absence of metacognitive deficit in schizophrenia) versus H1 (presence of metacognitive deficit in schizophrenia) by computing the ratio of the marginal likelihoods of each hypothesis (i.e., a Bayes factor or $BF_{10}$). We used the interpretation of $BF_{10}$ given by Wagenmakers et al. (2018), which translates continuous $BF_{10}$ values into a categorical scheme. Thus, we considered the relative strength of evidence for hypothesis H1 over H0, to be anecdotal if the $BF_{10}$ was $\in$ [1, 3], moderate if $\in$ [3, 10], strong if $\in$ [10, 30], very strong if $\in$ [30, 100] and extremely strong if > 100.

For subgroup analyses, we retrieved the statistics (mean and 95% CrI) of the difference between the two posterior distributions obtained in each group. Then we assessed in each case under which hypothesis (H0: absence of deficit or H1: existence of a deficit) the data was the most plausible.To assess the extent to which metacognitive performance was contaminated by first-order performance in studies which did not control for it, we fitted a meta-regression model identical to M1 with the z-scores of first-order performance as an additional continuous regressor. Based on the literature (Faivre et al., 2020), we specified an informative Gaussian prior (m = 0.56, sd = 0.24) for the slope of the meta-regression.

Risk of bias: The risk of bias regarding selection, comparability and outcome was assessed in parallel by two raters (MR and PS) and intraclass correlation (ICC) scores of agreement were computed with the R package irr (Gamer et al., 2012)

### Supplementary results

We assessed whether our selection of studies contained any extreme effect size values via a leave-one-out sensitivity analysis, which computes the effect sizes for each fold of n-1 studies, with n the total number of selected studies. This analysis revealed a strongly deviant study driving the overall effect size 4 standard deviations above the mean (Fig S1). This study was therefore excluded.

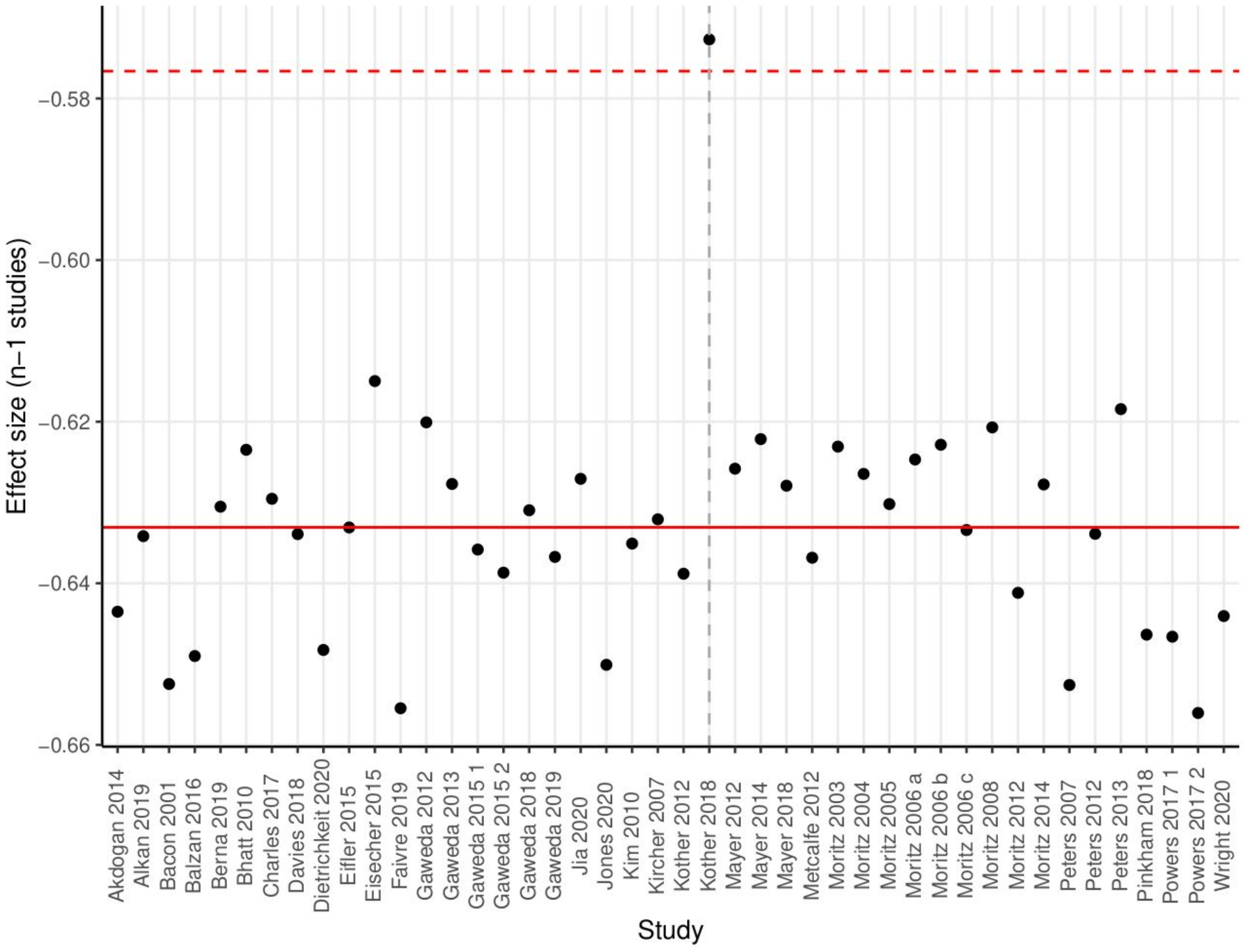


Figure S1: Effect size for each fold of n-1 studies. The horizontal red solid line indicates the average effect size, the dashed red line is four standard deviations above the mean. The vertical grey dashed line points to the deviant article, which has been excluded from our analysis.

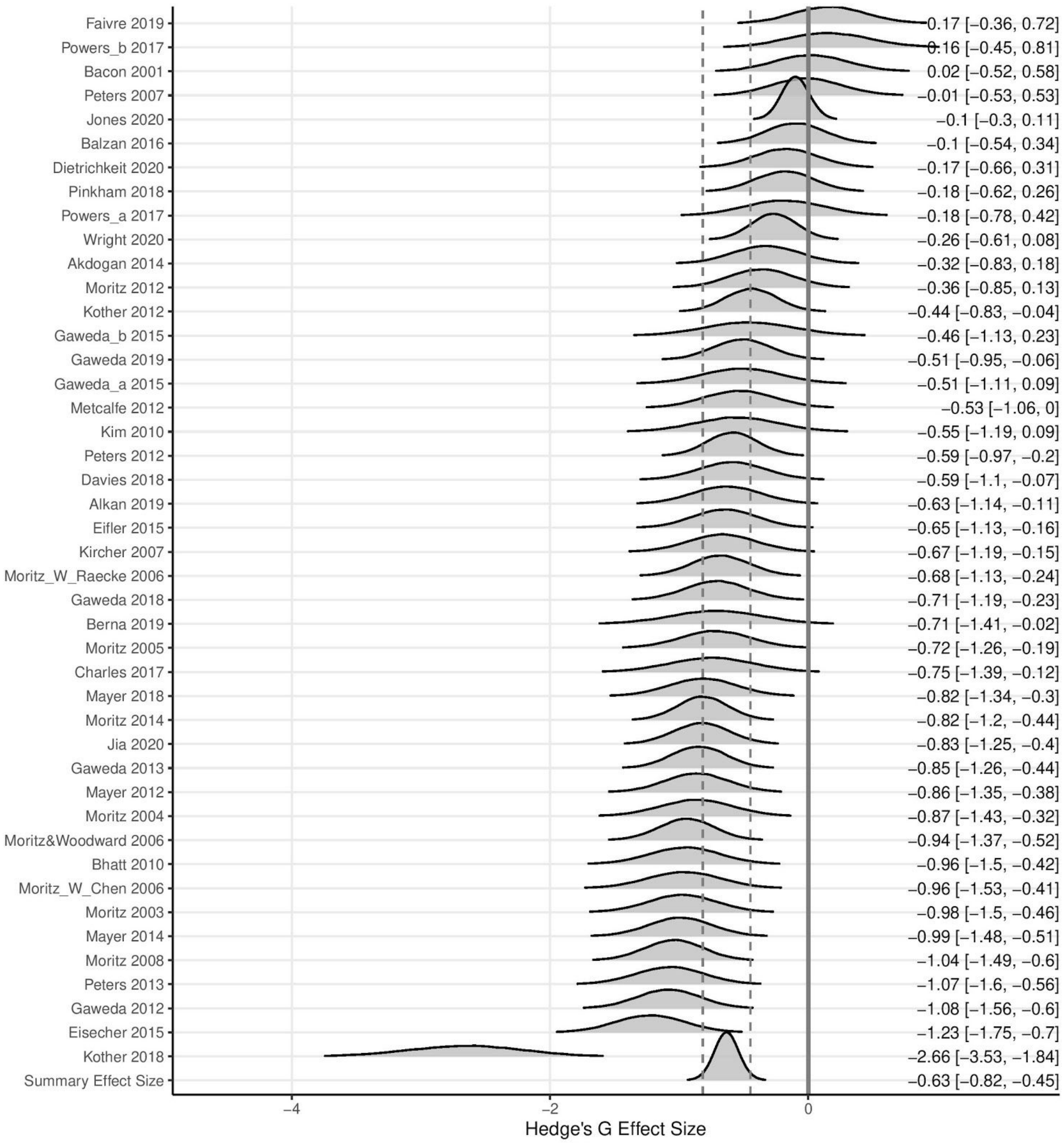


Figure S2: Forest plot of the metacognitive deficit in schizophrenia including the outlier study. Left: Authors with publication year; Middle: posterior distribution of the effect size; Right: mean and 95% CrI of the posterior distribution. The summary effect size is displayed on the last row: the solid vertical grey line is centred on zero (i.e., equivalent metacognitive performance between groups), and the dashed vertical lines depict the boundaries of the 95% CrI.

### Robustness analysis

To assess the influence of our choice of prior on effect size estimates, we re-ran the model M1 with a set of different priors $\alpha$ varying in mean and SD (Fig.S3). Except for very informative priors (SD = 0.1), results were robust to prior variations, in support of our main findings.

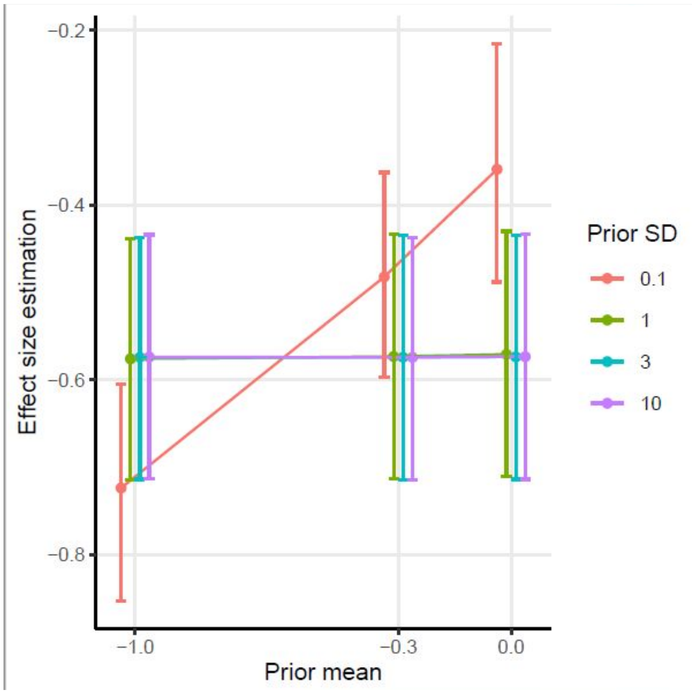


Figure S3: Metacognitive deficit effect size estimated by M1 with priors $\alpha$ varying in means (x-axis) and SDs (0.1: red; 1: green; 3: cyan; 10: purple).

The analysis of the moderating role of first-order performance was motivated by a significant moderation factor (Qbetween = 6.07, df = 1, p = .014), which means that the effect size was related to the control of first-order performance. Regarding subgroup analyses, $I^2$ for the non-controlled and controlled sub-groups were 0.65 and 0.65, corresponding to a reduction of 1.52% compared to $I^2$ across all studies. Q-statistic remained significant for the non-controlled and controlled sub-groups. We assessed the influence of performance-matching with a model identical to M1 including performance-matching as an additional binary predictor, with a Gaussian prior centered on -0.3 (SD = 1).

To examine the correlation between cognitive and metacognitive deficits among studies which did not control for first-order performance between groups, we performed a meta-regression by adding the standardized cognitive deficit as a continuous predictor to the model M1. On the basis of a previous study (Faivre et al., 2020), we specified a Normal prior with mean = 0.56 and SD = 0.24 for the slope parameter (the value of 0.24 corresponded to 0.1 before standardization). The mean slope value was b = 0.15, 95% CrI [-0.003, 0.30], with 97.3% of the slope estimates above 0, and very strong evidence in support of our hypothesis for a positive relationship between cognitive and metacognitive deficits ($BF_{10}$ = 36). Although a prior with SD = 0.24 is quite informative, the robustness analysis revealed stable patterns for a prior with mean = 0.56 (Fig.S5).

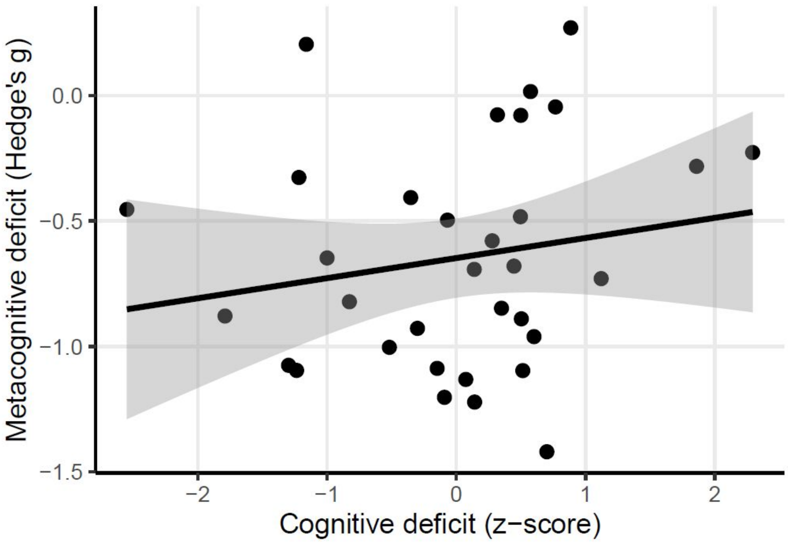


Figure S4: Meta-regression of the metacognitive deficit as a function of the cognitive deficit for studies which do not control for first-order performance. Each data point corresponds to one study (N = 33).

### Robustness analysis

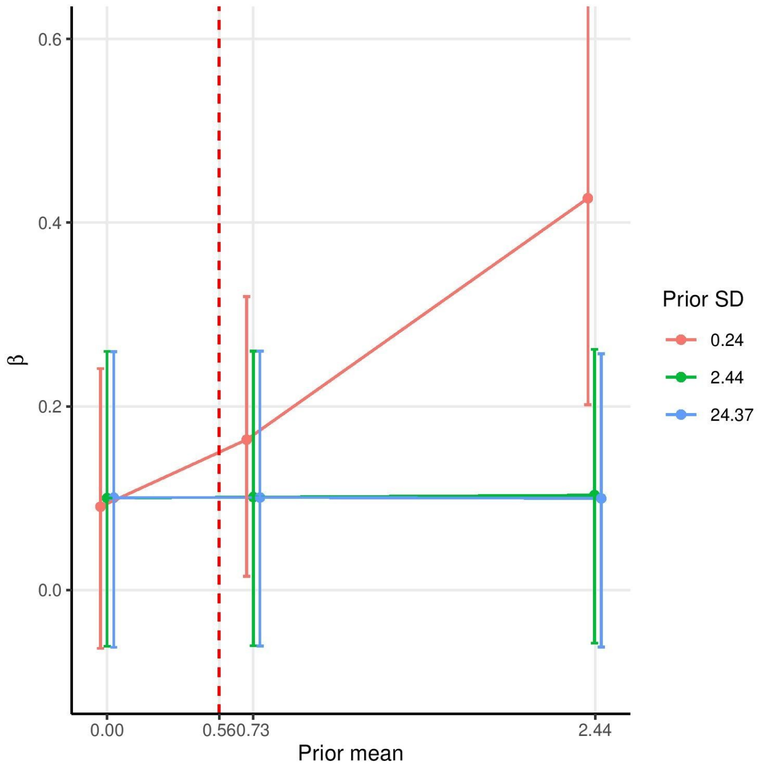


Figure S5: Slope estimates for the relationship between cognitive and metacognitive deficits under priors varying in means (x-axis, 0.73 and 2.44 corresponding to 0.3 and 1, respectively, after standardisation) and SDs (0.24: red; 2.44: green; 24.37: blue, corresponding to 0.1, 1, 10, respectively, after standardisation). The vertical red dashed line indicates the prior's mean value specified in our analysis.

Regarding metacognitive deficits across cognitive domains, $I^2$ for the memory, perception and other domain sub-groups were 0.52, 0.64 and 0 respectively, corresponding to a reduction of 21%, 3%, and 100% compared to the global analysis. Q-statistic remained significant for the memory and perception sub-groups. We performed a sub-group analysis with a weakly informative prior with mean = -0.3, and SD = 1 for the effect of cognitive domains. We found extremely strong evidence supporting a greater influence of memory vs. perception studies on the metacognitive deficit (m = -0.38, 95% CrI [-0.66, -0.09], $BF_{10}$ = 203). This pattern was even more pronounced when memory was compared with other domains (social and agency; m = -0.49, 95% CrI [-0.83, -0.12], $BF_{10}$ = 182).

Finally, we performed meta-regressions between the metacognitive deficits and clinical variables, with a prior of mean 0 and SD = 1. We had pre-registered the hypothesis of a negative correlation between meta-performance and PANSS positive scores. However, meta-regression analyses provided inconclusive evidence regarding the influence of symptom severity on the metacognitive deficit: $BF_{01}$ = 1.14 for PANSS total scores (N = 35), $BF_{01}$ = 1.11 for PANSS positive scores (N = 33) and $BF_{01}$ = 1.38 for PANSS negative scores (N = 34). Similarly, we found no evidence for an association between metacognitive performance and pharmacological treatment (N = 21), with an evidence ratio ($BF_{01}$ = 1.01) suggesting inconclusive data (Fig. S6).

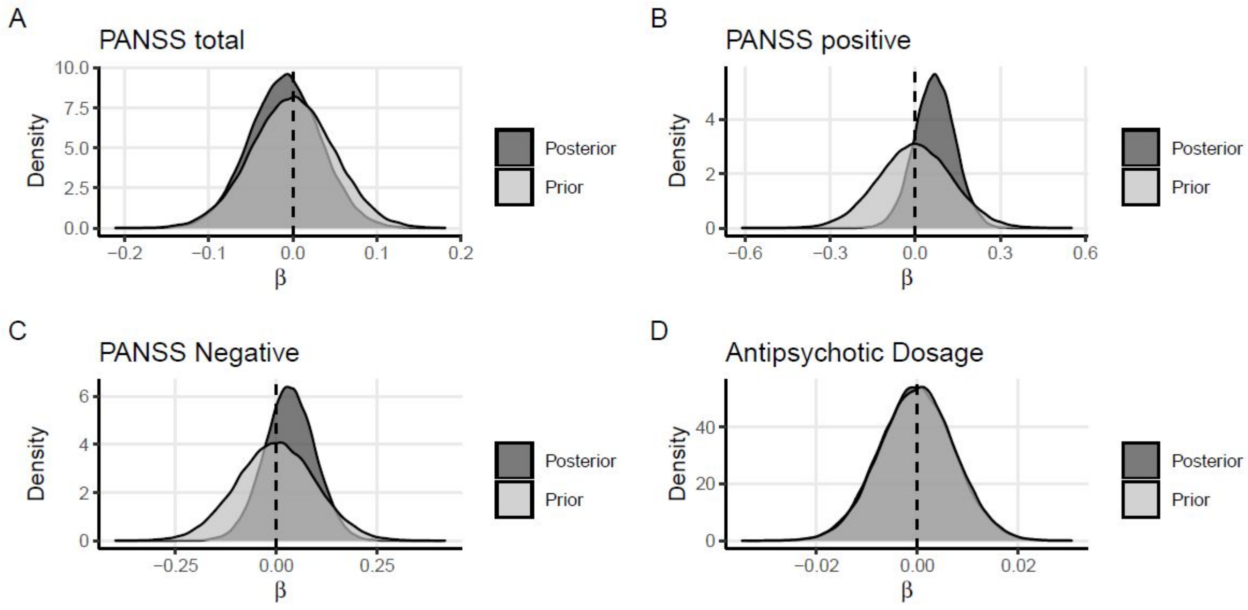


Figure S6: Meta-regressions of the metacognitive deficit with PANSS total scores (A), PANSS positive scores (B), PANSS negative scores (C), and antipsychotic dosage (D).The x-axes represent the posterior estimates for the slope parameter. Posterior and prior distributions are depicted in dark gray and light gray, respectively.

Similar results were found between first-order cognitive deficits and clinical features (Fig. S7) : $BF_{01}$ = 1.08 for PANSS total scores (N = 35), $BF_{01}$ = 1.34 for PANSS positive scores (N = 33), $BF_{01}$ = 1.35 for PANSS total scores (N = 34), $BF_{01}$ = 1.01 for PANSS total scores (N = 21).

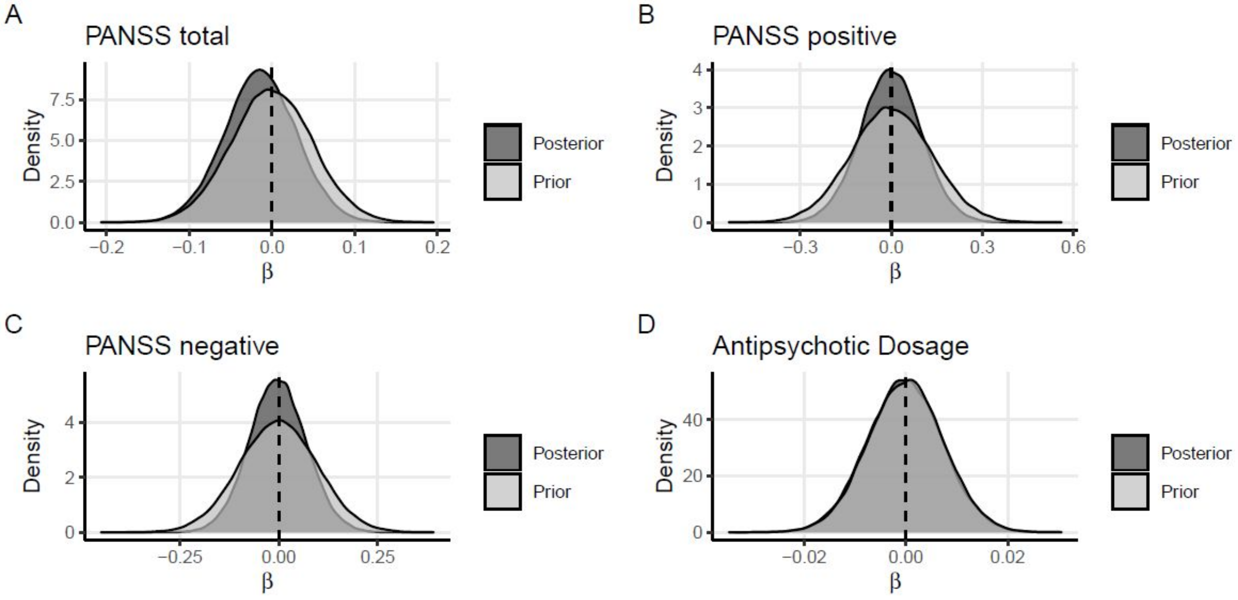


Figure S7: Meta-regressions of the cognitive deficit with PANSS total scores (A), PANSS positive scores (B), PANSS negative scores (C), and antipsychotic dosage (D). The x-axes represent the posterior estimates for the slope parameter. Posterior and prior distributions are depicted in dark gray and light gray, respectively.

The risk of bias was assessed using the Newcastle Ottawa Scale. The total ICC score (two way model, agreement type, single unit) revealed an average agreement between the two raters (MR and PS) of 0.55 according to interpretation schemes given by Koo & Li (2016). We then targeted the studies for which there were more than two divergences out of nine between the two raters. The six studies which reached this criterion were assessed again by two others raters (NF and PR). For these six studies, the final NOS score was obtained by averaging the scores given MR, PS, NF and PR.

About half of the studies included in this meta-analysis were rated as poor according to the interpretation scheme provided by the Newcastle Ottawa Scale (Fig. S8-9).

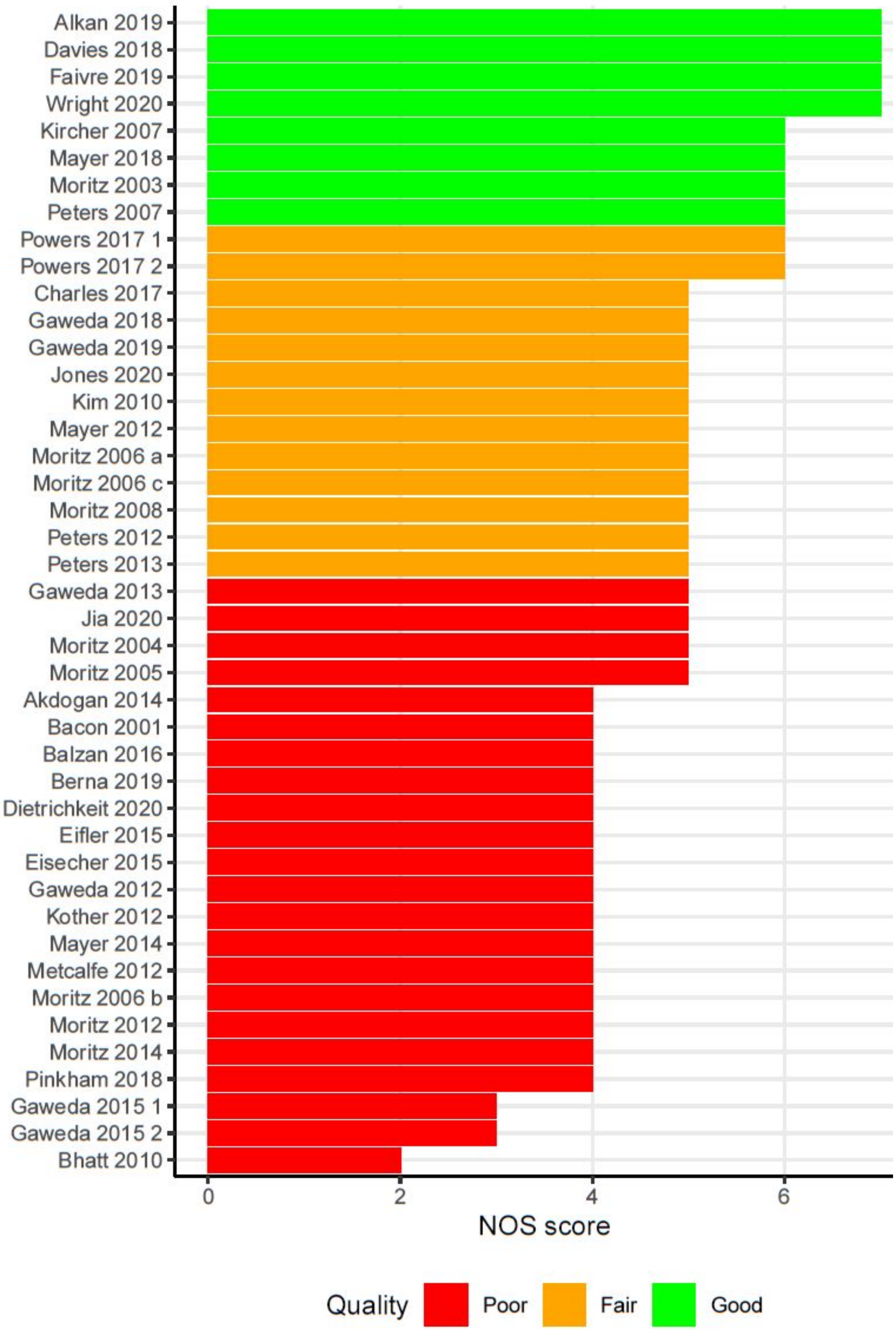


Figure S8: NOS quotation for each article included in the present meta-analysis

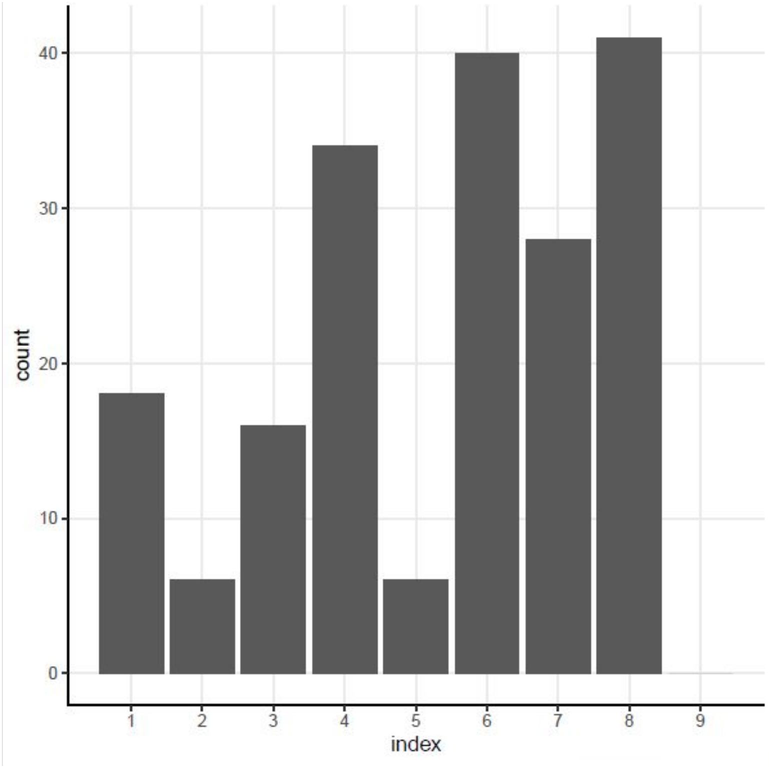


Figure S9 : The nine NOS items are presented on the x-axis: 1: Case definition adequacy, 2: Case representativeness, 3: Control selection, 4: Definition of controls, 5: Control for first-order performance between groups, 6: Control for Age/QI between groups , 7: Computerized protocol, 8: Same protocol for both groups, 9: Non-response rate. The y-axis represents the total number of articles which were granted a point for each NOS item.

| Study | PANSS (total) | PANSS (positive) | PANSS (negative) | Chlorpromazine (mg) | Manual |
|---|---|---|---|---|---|
| Akdogan 2014 | 69 ± 27.8 | 14.9 ± 4.6 | 18.8 ± 10.5 | Not reported | DSM4-TR |
| Alkan 2019 | Not reported | 5.24 ± NA | 5.76 ± NA | 264.8 ± NA | Not reported |
| Bacon 2001 | 67.62 ± 11.5 | 44.76 ± 16 | 54.48 ± 20.3 | 348 ± 273 | DSM3-R |
| Balzan 2016 | 11.08 ± 3.12 | Not reported | Not reported | Not reported | DSM4 |
| Berna 2019 | 75.13 ± 19.68 | 20.5 ± 5.04 | 17.88 ± 6.66 | Not reported | DSM4-TR |
| Bhatt 2010 | Not reported | Not reported | Not reported | Not reported | RDC |
| Charles 2017 | Not reported | 11.3 ± 3.4 | 15.5 ± 4.4 | 224.3 ± 103 | DSM4 |
| Davies 2018 | Not reported | 5.43 ± 2.44 | 5.4 ± 2.8 | 279.4 ± 150.2 | Not reported |
| Dietrichkeit 2020 | 51.69 ± 13.02 | Not reported | Not reported | Not reported | Not reported |
| Eifler 2015 | 67.41 ± 16.01 | 15.75 ± 5.34 | 16.59 ± 6.54 | 425.96 ± 183.68 | DSM4-R |
| Eisecher 2015 | 85.67 ± 18.62 | 22.95 ± 5.04 | 18.26 ± 7.02 | Not reported | DSM4 |
| Faivre 2019 | 78.5 ± 12.79 | 17.2 ± 5.85 | 20.5 ± 6.44 | 439.7 ± 27.08 | DSM5 |
| Gaweda 2012 | 72 ± 13.21 | 13.2 ± 5.36 | 19.54 ± 6.89 | Not reported | MINI |
| Gaweda 2013 | 62.57 ± 11.37 | 16.81 ± 5.43 | 17.81 ± 6.25 | Not reported | DSM4 |
| Gaweda 2018 | 87.64 ± 14.58 | 11.5 ± 4.3 | 3.72 ± 1.18 | 405.98 ± 175.04 | DSM4 |
| Gaweda 2019 | 46.81 ± 12.27 | 12 ± 4.9 | 13 ± 5.22 | Not reported | DSM5 |
| Gaweda_ 2015 | 66 ± 17.1 | 20.15 ± 8.07 | 18.23 ± 4.56 | Not reported | MINI |
| Gaweda_ 2015 | 79.55 ± 16 | 23.88 ± 6.97 | 19.33 ± 10.13 | Not reported | MINI |
| Jia 2020 | 54.6 ± 12.9 | 13.2 ± 5.6 | 14.1 ± 6.4 | 369 ± 397 | ICD-10 |
| Jones 2020 | Not reported | Not reported | Not reported | Not reported | DSM4 |
| Kim 2010 | 22.56 ± 6.6 | 20.4 ± 7.8 | 22.7 ± 10.1 | Not reported | DSM4 |
| Kircher 2007 | 66.4 ± 21.4 | 17.3 ± 7.2 | 17.2 ± 6.5 | 411 ± 257 | DSM4 |
| Kother 2012 | 58.28 ± 15.82 | 15.2 ± 8.21 | 14.14 ± 6.34 | Not reported | MINI |
| Mayer 2012 | 20.8 ± 5.74 | 24.17 ± 10.23 | 31.1 ± 13.81 | 361.12 ± 381.78 | DSM4 |
| Mayer 2014 | Not reported | 25.33 ± 11.12 | 32.53 ± 14.95 | 329.23 ± 299.36 | DSM4 |
| Mayer 2018 | 31.29 ± 7.92 | 14.92 ± 4.27 | 16.17 ± 6.31 | 501.13 ± 265.57 | DSM5 |
| Metcalfe 2012 | 46.1 ± 5.8 | Not reported | 36.89 ± 12.2 | Not reported | DSM4 |
| Moritz 2003 | 63.3 ± 12.3 | 8.86 ± 4.4 | 7.62 ± 3 | 253.04 ± 189.7 | DSM4 |
| Moritz 2004 | 13.84 ± 4.51 | Not reported | Not reported | 676.59 ± 523.86 | DSM4 |
| Moritz 2005 | 17.33 ± 7.35 | Not reported | Not reported | 671.6 ± 494.4 | DSM4 |
| Moritz 2008 | 62.24 ± 18.37 | Not reported | Not reported | Not reported | ICD-10 |
| Moritz 2012 | 60.04 ± 11.11 | Not reported | Not reported | Not reported | DSM4 |
| Moritz 2014 | Not reported | 1.8 ± 0.39 | 2.32 ± 0.54 | Not reported | Not reported |
| Moritz_a 2006 | 68.45 ± 17.48 | Not reported | Not reported | 687.88 ± 864.69 | DSM4 |
| Moritz_b 2006 | Not reported | 9.67 ± 4.2 | 12.77 ± 6.17 | Not reported | DSM4 |
| Moritz_c 2006 | 66.31 ± 16.62 | Not reported | Not reported | Not reported | DSM4 |
| Peters 2007 | 55.64 ± 8.52 | 9.17 ± 4.29 | 5.3 ± 2.12 | Not reported | DSM4 |
| Peters 2012 | 54.7 ± 13.27 | 14.81 ± 6.28 | 11.7 ± 4.94 | Not reported | DSM4 |
| Peters 2013 | 49.29 ± 9.75 | 12.66 ± 4.76 | 11.59 ± 4.63 | 368.87 ± 260.63 | MINI |
| Pinkham 2018 | 30.48 ± 6.44 | 17.54 ± 5.68 | 12.23 ± 3.2 | 452.26 ± 416.14 | MINI/SCID |
| Powers_ 2017 | 61.2 ± 7.94 | 19.67 ± 4.14 | 15.2 ± 4.8 | 431.46 ± 90.92 | DSM4 |
| Powers_ 2017 | 54.43 ± 10.33 | 14.93 ± 3.26 | 14.14 ± 4.19 | 330.43 ± 66.28 | DSM4 |
| Wright 2020 | Not reported | 12.4 ± 4.7 | 11.5 ± 4 | Not reported | Not reported |

Table S1: Summary of clinical data for each study (PANSS total, PANSS positive, PANSS negative, chlorpromazine equivalent and diagnostic tool).
